# Identifying drivers of pertussis disease resurgence pilot study: Final report

**DOI:** 10.1101/2025.05.22.25328117

**Authors:** Patricia Therese Campbell, Yoon Hong Choi, Manoj Gambhir, Jodie McVernon

## Abstract

Three separate transmission models were developed to investigate the drivers of pertussis resurgence in the United States (US), United Kingdom (UK) and Australia. While these models were all able to reproduce pertussis trends in the setting for which they were developed, they include different assumptions about infection, immunity and vaccination, reflecting the uncertainties of pertussis epidemiology. A key question is whether, despite these differences, these models produce consistent results across settings with diverse vaccination schedules, epidemiologic histories and surveillance systems. A pilot study was conducted in 2016 for the World Health Organization to examine whether findings from the US, UK and Australian models were generalisable to other country contexts. In Stage I of the study, each of the UK, US and Australian models was applied to the other two settings, using the previously identified best fitting parameter sets. The major aim of this phase was to determine whether there is overall evidence that the models may be applied to high-income countries with long histories of pertussis vaccination. In Stage II of the study, each of the UK, US and Australian models was fitted to the other two settings using the same model selection techniques originally applied, to evaluate consistency of model results and identify broadly congruent characteristics of pertussis transmission and immunity. A major aim of this phase was to determine whether, for a given country, the different models provided similar reasons for pertussis resurgence. Each of the models worked well in the setting for which they were designed, but were not able to capture all of the epidemiologic features in different countries. Limited availability of historical data on vaccination, age-specific burden of disease and the relationship between infection and disease is likely to make fitting of the models to other settings without adaptation challenging.

## 1 Background

Three separate transmission models have been developed to investigate the drivers of pertussis resurgence in the United States (US), United Kingdom (UK) and Australia. While these models have all been able to reproduce pertussis trends in the setting for which they were developed, they include different assumptions about infection, immunity and vaccination, reflecting the uncertainties of pertussis epidemiology. A key question is whether, despite these differences, these models produce consistent results across settings with diverse vaccination schedules, epidemiologic histories and surveillance systems.

A pilot study in two stages was conducted to examine whether findings from the US, UK and Australian models are generalisable to other country contexts:

- Stage I: Application of each of the UK, US and Australian models to the other two settings, using the previously identified best fitting parameter sets. The major aim was to determine whether there is overall evidence that the models may be applied to high-income countries with long histories of pertussis vaccination;
- Stage II: Fitting each of the UK, US and Australian models to the other two settings using the same model selection techniques originally applied, to evaluate consistency of model results and identify broadly congruent characteristics of pertussis transmission and immunity. A major aim was to determine whether, for a given country, the different models provide similar reasons for pertussis resurgence. The work done in this stage was also used to ascertain the feasibility of a similar, larger study across multiple settings with diverse vaccination and disease histories.

## 2 Brief model descriptions

All of the models are age-structured, compartmental deterministic models. Details of each model are provided below, including the methods used to fit the model to the country for which it was developed. A representation of the three models is included in figure 1.

**Figure 1:**
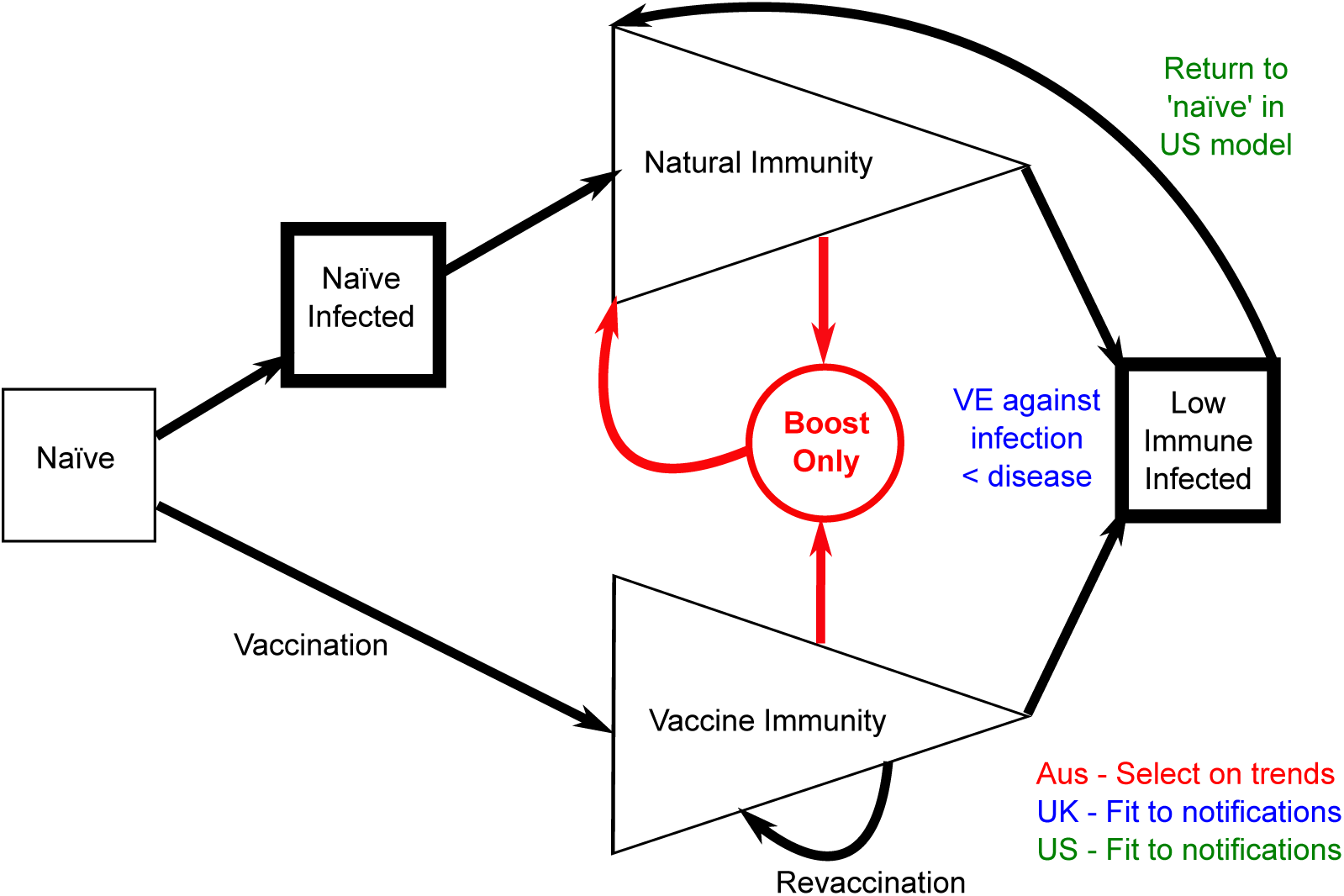
Representation of Australian, UK and US pertussis models. In the Australian model, immunity may be boosted without contributing to the force of infection, and *infections* in both the Näıve infected and Low immune infected compartments are reported. In the UK model, notifications are estimated as a proportion of infections in the Näıve infected compartment and a smaller proportion of infections in the Low immune infected compartment. In the US model, notifications are estimated as a proportion of infections in the Näıve infected compartment only, however, individuals with fully waned immunity return to the Näıve infected compartment.

### 2.1 Australia

The Australian model [1] separates the näıvely susceptible population from those whose immune system has been primed by pertussis infection or vaccination. Primed individuals are further distinguished by whether their most recent experience was infection or vaccination. As priming of the immune system is assumed to permanently modify infection risk and characteristics, primed individuals never return to the näıve susceptible state. Immunity against pertussis infection is determined by whether the previous experience was vaccination or infection, and the length of time that has elapsed since the last infection or vaccination. The model’s parameters include terms to quantify the duration and action of immunity, duration and degree of infectiousness following infection, vaccine coverage and efficacy and mixing patterns within the population. Parameter selection used a filtering process, whereby 200,000 simulations of the model were run using alternative parameter sets and those that reproduced the broad epidemiologic trends in Australia (2,321 parameter combinations) were retained for further analysis.

### 2.2 United Kingdom

The UK model [2] distinguishes sub-populations with vaccine derived and infection induced immunity, as well as protection against clinical disease and infection. While infection induced immunity is assumed to provide complete protection against both infection and disease for a period of time, vaccination fully protects against notifiable disease, but possibly provides only partial protection against infection and transmission. Infections in näıve individuals are assumed to be more likely to be notified than subsequent infections. As with the Australian model, following vaccination or infection there is no return to the näıve susceptible state. Once immunity has waned, primed individuals become infected at the same rate as näıve susceptibles. This model incorporates a parameter to quantify the proportion of new infections that develop notifiable symptoms. The changing age distribution of the UK population was included in the model. A range of plausible values for each parameter were chosen and each combination of these parameter values simulated to estimate those combinations producing the best fit to age-specific disease notifications in the UK. The 2,000 best fitting combinations were retained and these combinations used for further analysis.

### 2.3 United States

The US model [3] separates the population with waned vaccine immunity from the population with waned infection-derived immunity. Unlike the Australian and UK models, individuals with waned infection-derived immunity are not distinguished from those experiencing their first infection. Only infections in the primary-infected compartment, which occur in näıvely susceptible individuals and those with waned immunity (both infection-derived and vaccine) are considered to generate disease. That is, while infections can occur in vaccinated individuals whose immunity has not yet waned, these infections are not considered to be reported, although they are infectious to some extent. Like the UK model, the changing age distribution of the US population was included. A series of eight models were fitted in a Bayesian framework to annual US incidence data to identify parameter combinations that fit the data, with candidate models compared using the Deviance Information Criterion. The fitting process identified a model with essentially lifelong whole-cell vaccine- and infection-derived immunity as the best fitting model.

## 3 Stage I: Application of existing models and parameter estimates to other high income settings

In this stage of the project, each model was simulated with the previously identified best-fit parameters, applying the vaccination schedule and coverage for the US, UK and Australia.

### 3.1 Population assumptions

To run each of the three models with the previously identified parameter sets requires information on population size for a single year (Australian model) or yearly population distribution (UK and US models). As the Australian model assumes a flat population distribution and applies the population size in 2010 for the period over which the model is run, implementing a change of population size to reflect the UK and US populations was a trivial matter and so this change was made for this stage. To apply a *different* changing population size to the UK and US models would have involved re-fitting the model to generate the correct population mixing patterns. Given the similarities between these three high income countries (increasing population, ageing population structure, post-war population boom), as a simplification for this stage of the project, the demographics of the country for which the model was developed were used to run the simulations (UK and US models). Results presented for this stage of the project are defined as:

- Australian model: The annual number of low immunity infections (occurring from either the näıve susceptible state or the fully waned state) per 100,000 population;
- UK model: The annual number of notifications occurring in a population with the demographics of the UK; and
- US model: The annual number of infections per 100,000 occurring in a population with the demographics of the US.

### 3.2 Vaccination assumptions

Other than population data, the only other data required by the models are the applicable vaccination coverage and schedules, with varying availability:

- Australia: High quality coverage data for children under 7 years of age was available from 1996. Prior to that time, a number of different methodologies were used to estimate coverage with limited reliability, while changes to the vaccination schedule are reasonably well documented;
- UK: Vaccination coverage data were available for almost the entire period, with estimation required between 1957 and 1966, assumed to be the same as the 1967 published rate; and
- US: Vaccination coverage for the primary course is available from 1962, and for the 18 month booster from 1995. No definitive dates of implementation for the 18 month and 4–5 year old booster were found, nor any coverage data for the 4–5 year booster, although implementation of these doses appears to have occurred quite early [4]. Adolescent coverage was available from the date of implementation in 2006.

### 3.3 Comparison of model results for each setting

Each of the three models was run using the vaccination schedules for each of the three countries. For comparison purposes, the results of these simulations are presented for the Australian (figure 2), UK (figure 3) and US (figure 4) vaccination schedules. Thus each set of three subfigures shows the behaviour of each of the three models when the same vaccination schedule and coverage is applied.

**Figure 2:**
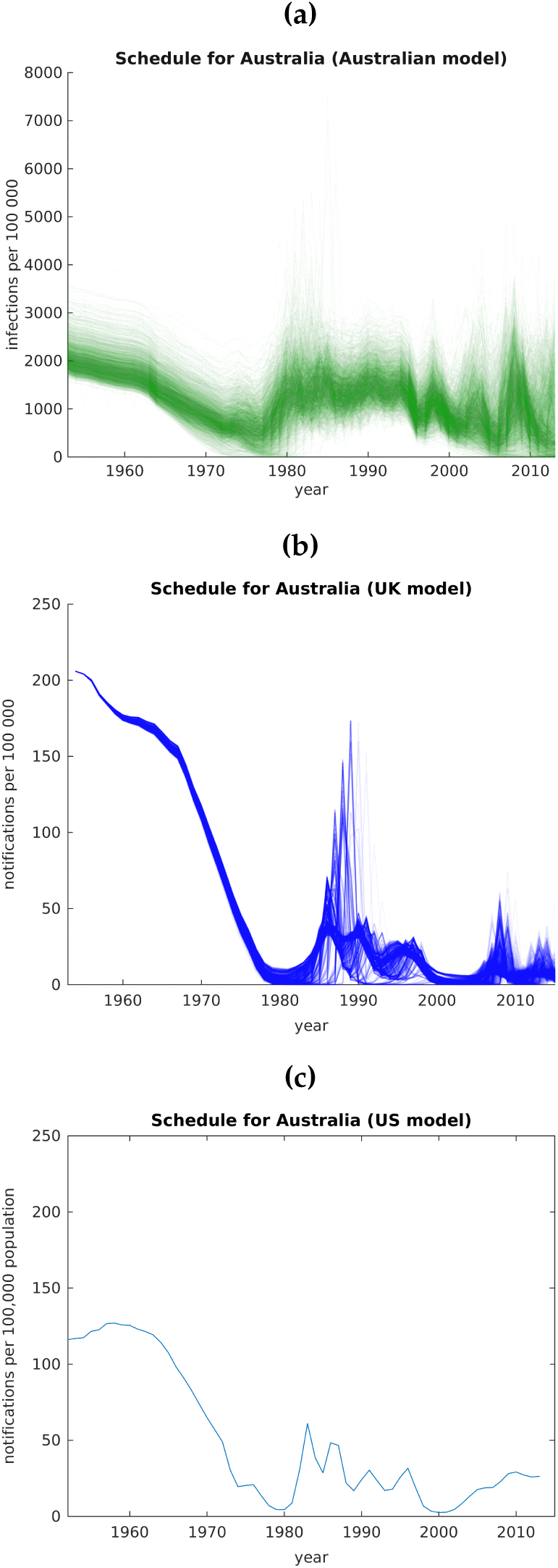
Australian pertussis epidemiology in a) the Australian model, b) the UK model, c) the US model. Incidence was generated by simulating each of the three models using existing ‘bestfit’ parameters and with the Australian vaccination schedule in place. Note that the Australian model produces infections per 100,000 (shown in green) while the UK and US models produce notifications (shown in blue).

**Figure 3:**
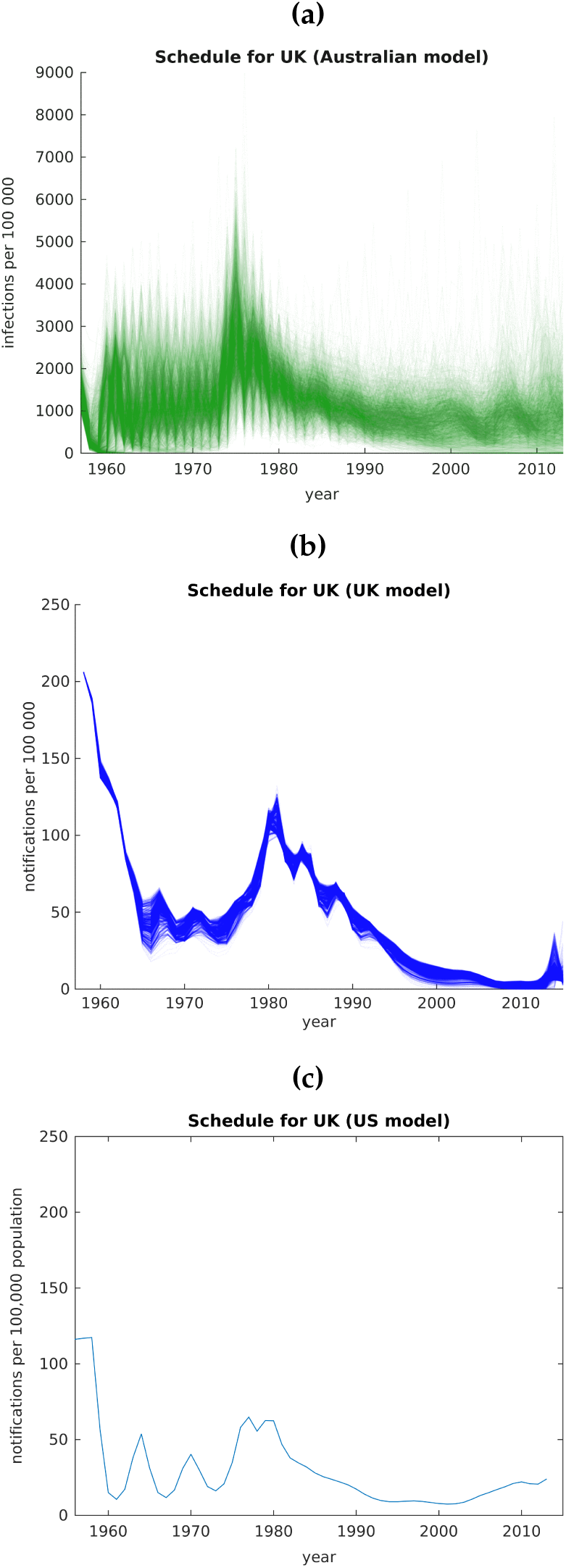
UK pertussis epidemiology in a) the Australian model, b) the UK model, c) the US model. Incidence was generated by simulating each of the three models using existing ‘bestfit’ parameters with the UK vaccination schedule in place. Note that the Australian model produces infections per 100,000 (shown in green) while the UK and US models produce notifications (shown in blue).

**Figure 4:**
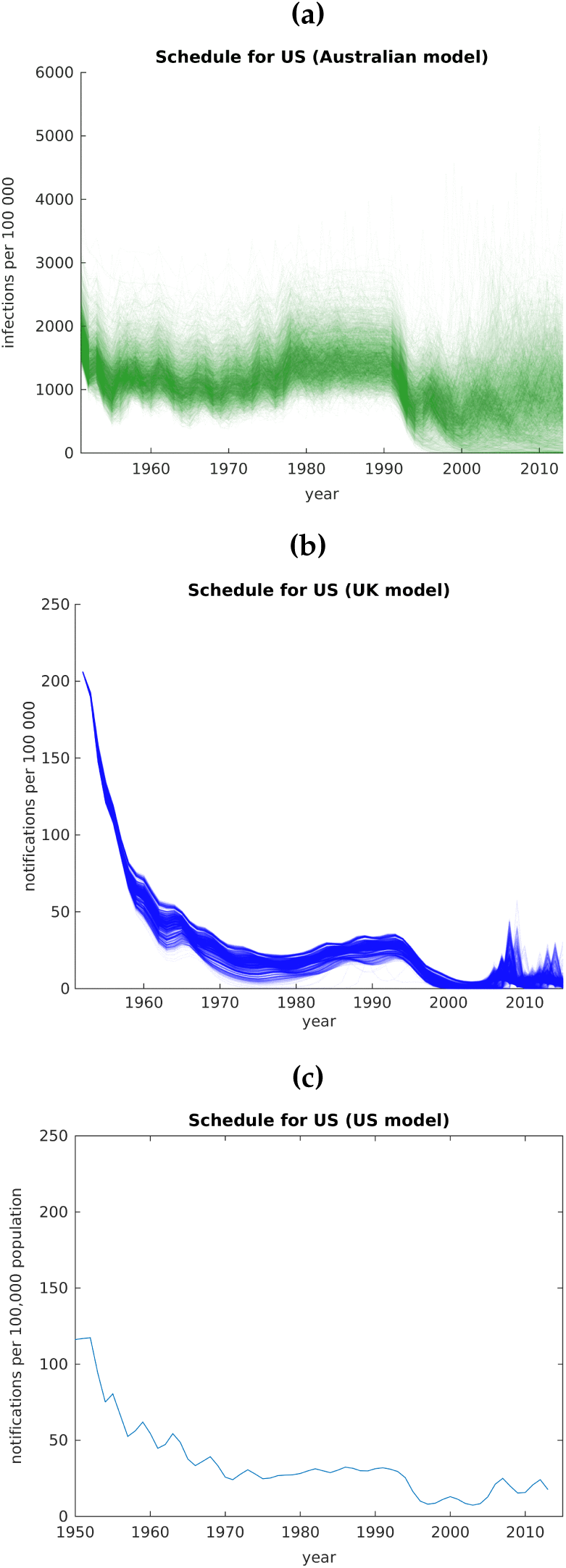
US pertussis epidemiology in a) the Australian model, b) the UK model, c) the US model. Incidence was generated by simulating each of the three models using existing ‘bestfit’ parameters with the US vaccination schedule in place. Note that the Australian model produces infections per 100,000 (shown in green) while the UK and US models produce notifications (shown in blue).

Generally, all models capture the decreases in incidence (either infections or notifications) when vaccination was first implemented, as well as subsequent rebounds when vaccination coverage fell away in the late 1970s and early 1980s. In particular, the timing of increases and decreases is reasonably consistent between all three models in each setting. In the US model, as soon as acellular vaccination is introduced for the primary course in any setting, incidence gradually rises, with the effect not as strong in the other models. Removing the 18 month booster from the Australian schedule produced a larger, more sudden rebound in both the Australian and the UK models than in the US model.

#### 3.3.1 Australian vaccination schedule

In figure 2, the Australian (a) and US (c) models represent infections per 100,000 population, while the UK (b) model produces notifications per 100,000 population. All three models capture the decrease in infections/notifications from the introduction of vaccination through to the late 1970s, then a resurgence through the 1980s when vaccination coverage fell and the 18 month booster was removed. The models generate a period of sustained activity through the 1990s, followed by a reduction in incidence in the late 1990s and early 2000s. All three models produce a large epidemic around 2008–2010, although the rise in the US model is a somewhat more steady increase than the Australian and the US models produce.

#### 3.3.2 UK vaccination schedule

In figure 3, the UK (b) model produces a reduction in notifications per 100,000 to around 25 % of the pre vaccination value through the late 1960s and early 1970s. The reduction in infections observed in the Australian (a) and US (c) models (which report infections per 100,000) when vaccination is implemented is not as large as the reduction reported in the UK model, which reports notifications. Oscillations in incidence are evident in all three models following the introduction of vaccination. All models capture the timing of the large resurgence in the late 1970s and early 1980s due to substantial falls in vaccination coverage, followed by a gradual decline as coverage returned to high levels. A slight increase in infections is observed in the Australian model through the 2000s, while the increase in the UK and US models is more marked around 2010. The reduced ability of the Australian model to generate large epidemics due to the switch to acellular vaccines is likely due to the large influence the removal of the 18 month booster had on parameter values for the Australian model. A notable trend in each of the three models is the delayed time to resurgence in the modelled UK results compared to Australia and the US.

#### 3.3.3 US vaccination schedule

In figure 4, each of the three models produces a similar pattern of infections/notifications, with a reduction upon introduction of vaccination followed by a gradual increase starting from the mid 1970s until the early 1990s. The UK (b) and US (c) models produce a defined epidemic around 2008–2010. Some of the parameter combinations for the Australian model also produce such an epidemic, but as mentioned previously, the parameter selection process for the Australian model was unable to determine a difference between whole cell and acellular vaccines based on the Australian experience, and so this epidemic is not as well defined in the simulations.

### 3.4 Summary

Running each of the three models in each of the three settings produced broadly concordant results despite the differences in demographic assumptions and the different model structures. This stage of the project provided evidence that the Australian, UK and US models may be applicable to high income countries with long standing vaccination programs.

## 4 Stage II: Fitting the Australian, UK and US models to other high income settings

In this stage of the project, the aim was to fit each of the three models to the other two settings, to determine new ‘best-fit’ parameter values for each model/setting combination.

### 4.1 Fitting procedures

Both the Australian and UK models use a filtering design, whereby a finite number of parameter combinations are pre-determined, and the model simulated with these parameter combinations followed by selection of the best fitting simulations. The US model uses Markov Chain Monte Carlo methods to fit incidence data, in which parameter space is searched for combinations that provide the best fit to the data.

#### 4.1.1 Fitting the Australian model

Latin Hypercube Sampling was used to sample parameters from plausible ranges and the Australian model then simulated with these parameter combinations (100,000 in total) and the vaccination schedule and coverage applicable to each country. Results were then filtered to match robust features of the observed epidemiology. When this model was originally run using Australian conditions, the removal of the 18 month booster dose from the Australian schedule exerted a large influence on epidemiology, and thus the criteria used to select parameter combinations may have contributed to the inability to distinguish a difference in immune characteristics induced by whole cell and acellular vaccines. In our original work on this model, we filtered on the condition of resurgence following withdrawal of the 18 month booster dose. This fourth criterion has been replaced in the list below^∗^.

For each country, key epidemiologic characteristics were extracted for each simulation, and simulations that reproduced all of the following criteria were retained:

- Australia:

– a pre-vaccination inter-epidemic period of between 2 and 4.5 years;
– a minimum four-fold reduction in infant (*<* 6 mth) infections following vaccine introduction;
– the adult population reaching high levels of seropositivity (80 %) at least once in the ten years from 1997; and
– low immunity infections from 2010–2012 monotonically increasing with each year of age for 5–9 year olds^∗^.
- UK:

– a pre-vaccination inter-epidemic period of between 2 and 4.5 years;
– a minimum four-fold reduction in infant (*<* 6 mth) infections following vaccine introduction;
– low immunity infections in 10–14 year olds occurring at a rate at least 1.3 times that of the total population (over 2010–2014); and
– maximum annual number of low immunity infections in the period 1990–2005 no more than one-quarter the number of low immunity infections occurring in the year before vaccination.
- US:

– a pre-vaccination inter-epidemic period of between 2 and 4.5 years;
– a minimum four-fold reduction in infant (*<* 6 mth) infections following vaccine introduction;
– low immunity infections from 2010–2012 monotonically increasing with each year of age for 5–9 year olds; and
– low immunity infections in 7–10 year olds occurring at a rate at least 4 times that of the total population (in 2008).

In the original application of the Australian model to the Australian population, four different contact matrices were evenly sampled. Based on the findings of the original modelling study, a single contact matrix was used for this pilot study, the POLYMOD ‘All contacts’ matrix for Great Britain [5].

#### 4.1.2 Fitting the UK model

Fitting of the UK model occurred in three steps:

- Step 1 - Pre-vaccination equilibrium:

– The pre-vaccination force of infection (FOI) and primary infection notification probability were generated for 45 combinations of the duration of natural immunity (9 values) and secondary infection reporting probability (5 values; relative to primary infections) by fitting to age-stratified pre-vaccination notification rates;
– No pre-vaccination age-stratified notification data are available for Australia. The population pre-vaccination notification rate was estimated at 250 per 100,000 population, based on the smoothed average over the period 1920–1940 before vaccines were available [6]. In the absence of any other information, it was assumed that the age distribution of cases in Australia was the same as in the UK, as the age distributions of the two populations were quite similar.
– Similarly, no pre-vaccination age-stratified notification data are available for the US. The population pre-vaccination notification rate was estimated at 150 per 100,000 population, based on published data [7, 3]. As was done for Australia, it was assumed that the age distribution of cases in the US was the same as for the UK.
- Step 2 - Fitting of whole cell vaccine era:

– Combinations of four model parameters (duration of natural immunity (9 values); secondary infection reporting probability (5 values); duration of whole cell vaccine (wP) immunity (11 values); degree of wP vaccine protection against infection (9 values)) were simulated using the model and the FOI sets estimated in step 1, for the 3,555 combinations with wP duration not greater than natural duration;
– Model generated notifications were compared to published notification data for the whole cell era, with Poisson deviance calculated. The parameter combinations corresponding to the best-fitting 5% of simulations were retained for fitting to the acellular data;
– For Australia, notification data are only available from 1991, and acellular vaccines were introduced from 1997–1999. Therefore age-stratified notifications in 5-yearly age blocks for the period 1993–1997 inclusive were used to fit model generated notifications in the whole cell era;
– For the US, age stratified data are not readily available and so total notifications for the period 1951 to 1991 were used to fit model generated total notifications in the whole cell era.
- Step 3 - Fitting of acellular vaccine era:

– Using the best-fitting parameter combinations identified in step 2, the model was run from the introduction of acellular vaccines to 2014, with the inclusion of two new parameters: duration of acellular vaccine (aP) immunity (11 values); and degree of aP vaccine protection against infection (9 values), on the proviso that aP duration was not greater than wP duration.
– Model generated notifications were compared to published notification data for the acellular era, with Poisson deviance calculated. The parameter combinations corresponding to the best-fitting 5% of simulations were retained for further analysis;
– For Australia, age-stratified notifications in 5-yearly age blocks for the period 2000– 2014 inclusive were used to fit model generated notifications in the acellular era;
– For the US, total notifications for the period 2000–2014 were used to fit model generated total notifications in the acellular era.

#### 4.1.3 Fitting the US model

Fitting of the US model to Australian and UK notification data was undertaken using Markov Chain Monte Carlo (MCMC) methods as described in the published study [3]. The MCMC process returns probability distributions for each parameter. Beginning with an initial starting value for each of the parameters to be fitted, the model is simulated and the likelihood that this simulation could give rise to the observed data is calculated from the model incidence. Next, a move from the current position in parameter space to a new position is proposed, with the standard deviation of the prior distribution for each parameter determining how far away the proposed position is in parameter space. If the (model) likelihood for this proposal is greater than for the current location, the proposal is accepted. Otherwise, the proposal is accepted probabilistically; proposals with substantially lower likelihoods are not likely to be accepted.

There were ten parameters to be fitted through the MCMC process, namely:

- transmission parameter;
- vaccine efficacy of whole cell vaccine;
- vaccine efficacy of acellular vaccine;
- duration of protection following natural immunity;
- duration of protection following whole cell vaccination;
- duration of protection following acellular vaccination;
- relative rate of mild infections compared to severe infections;
- relative infectiousness of mild infections compared to severe infections;
- two parameters describing the distribution of the reporting probability.

Based on advice from the model authors that age-stratified data can be extremely difficult to fit, the model was fit to overall population notification data. For the UK, records were available from 1956 to 2014, and for Australia from 1991 to 2014 (only 1993 to 2014 were used, due to known issues with the first few years of mandatory reporting).

A number of difficulties arose in using MCMC to fit the US model to UK and Australian data, primarily based around the choice of starting point in parameter space. Three approaches to selecting starting points for the MCMC process were tried:

1. using the starting values originally applied in the implementation of the US model to the US population:

- analysis of the MCMC results for Australia and the UK indicated a high level of similarity (’autocorrelation’) of accepted values for each parameter, meaning that from 30,000 simulations, only around 10 different values of each parameter were simulated, suggesting that a very large number of simulations would be required to provide good coverage of parameter space.
2. using starting values uniformly sampled from within the allowable range for each parameter:

- analysis of the MCMC results for Australia and the UK indicated that for each of the randomly selected initial starting values, which were generally not close to the expected mean of the parameter value, the fitting process had become ‘stuck’ at an extreme of the parameter distribution. Greater than 99% of the parameter proposals were rejected because each time a new value was proposed, it was outside the allowable limits for the parameter and therefore there was poor exploration of the parameter space.
3. using starting values randomly selected from the prior distribution for each parameter:

- five runs of 50,000 simulations each, for both the UK and Australia, were run to determine whether similar posterior parameter distributions could be obtained from different starting points. Selecting from the prior distribution was a stronger assumption than selecting uniformly across the range. The results produced using this method are presented in this report.

For each of the five runs, an unbiased sample of 200 parameter combinations was selected and the model was simulated with these parameters. Incidence patterns for one of these runs are presented in section 4.2.1, with those for the remaining four runs provided in the Appendix.

### 4.2 Results

#### 4.2.1 Incidence patterns

Each of the Australian, UK and US models produce similar behaviour for each of the three countries, although there are some differences in the ability of the models to reproduce epidemiologic characteristics (figures 5, 6 and 7). Common to all models and settings was a sensitivity to vaccination coverage, with the decline in incidence following the introduction of mass vaccination and subsequent rebound due to reductions in coverage captured well. Resurgence in each country in the acellular vaccine era was not reproduced to the same extent as resurgence due to falls in vaccination coverage.

**Figure 5:**
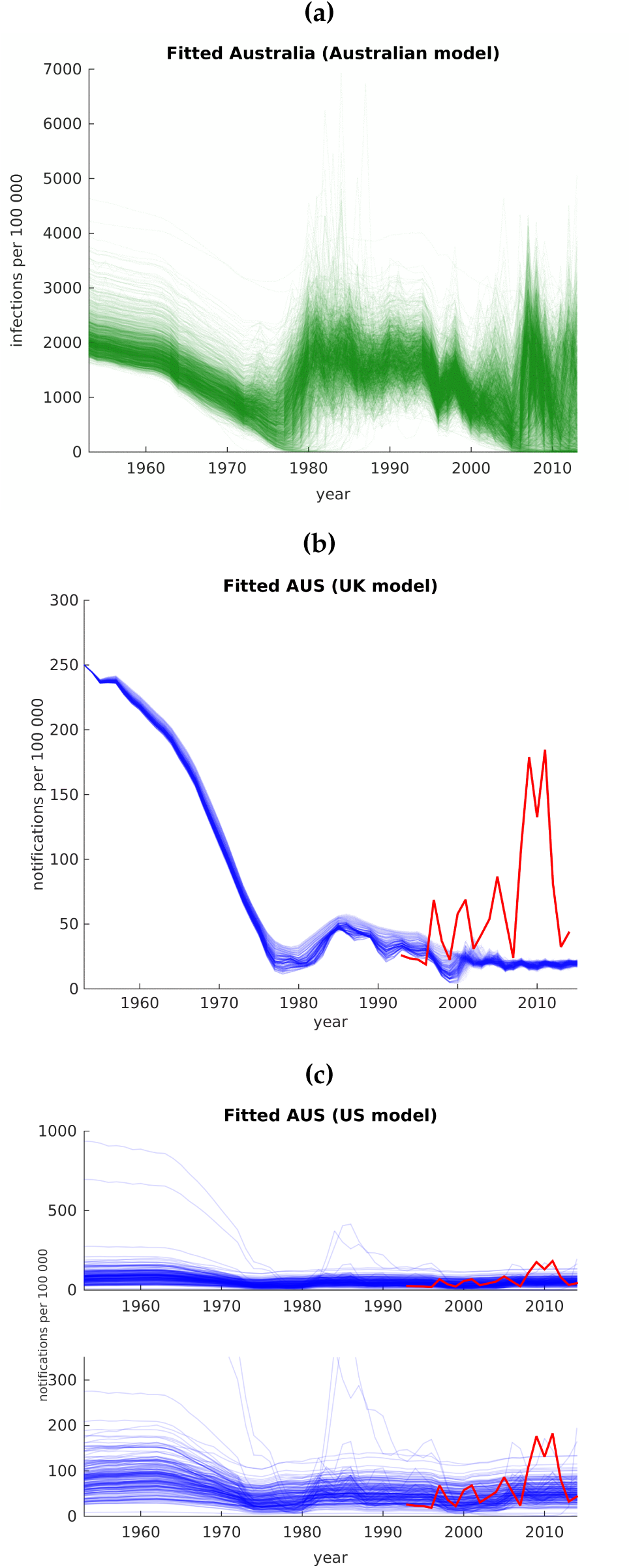
Fitted Australian pertussis epidemiology in a) the Australian model, b) the UK model, and c) the US model. Note that the Australian model produces infections per 100,000 (shown in green) while the UK and US models produce notifications (shown in blue). In b) and c), the red solid line shows Australia’s published notifications used to fit the models. Part c) shows simulations with the US model using two different scales to include all points — the bottom panel zooms in over the scale of the majority of simulations. All models produce the initial decline in incidence upon the introduction of mass vaccination, followed by an upsurge through the early 1980s as coverage fell and the 18 month booster was removed from the schedule for the first time. The relative magnitude of the increase through the 1980s was much greater in the Australian and US models than seen in the UK model. The Australian model captures the relative size of the 2008– 2012 epidemic following removal of the 18 month booster for the second time, a feature not captured by the UK model. An increase in notifications starting in 2005 can be seen in some of the US model simulations, returning to levels close to those produced by the model in the early days of vaccination.

**Figure 6:**
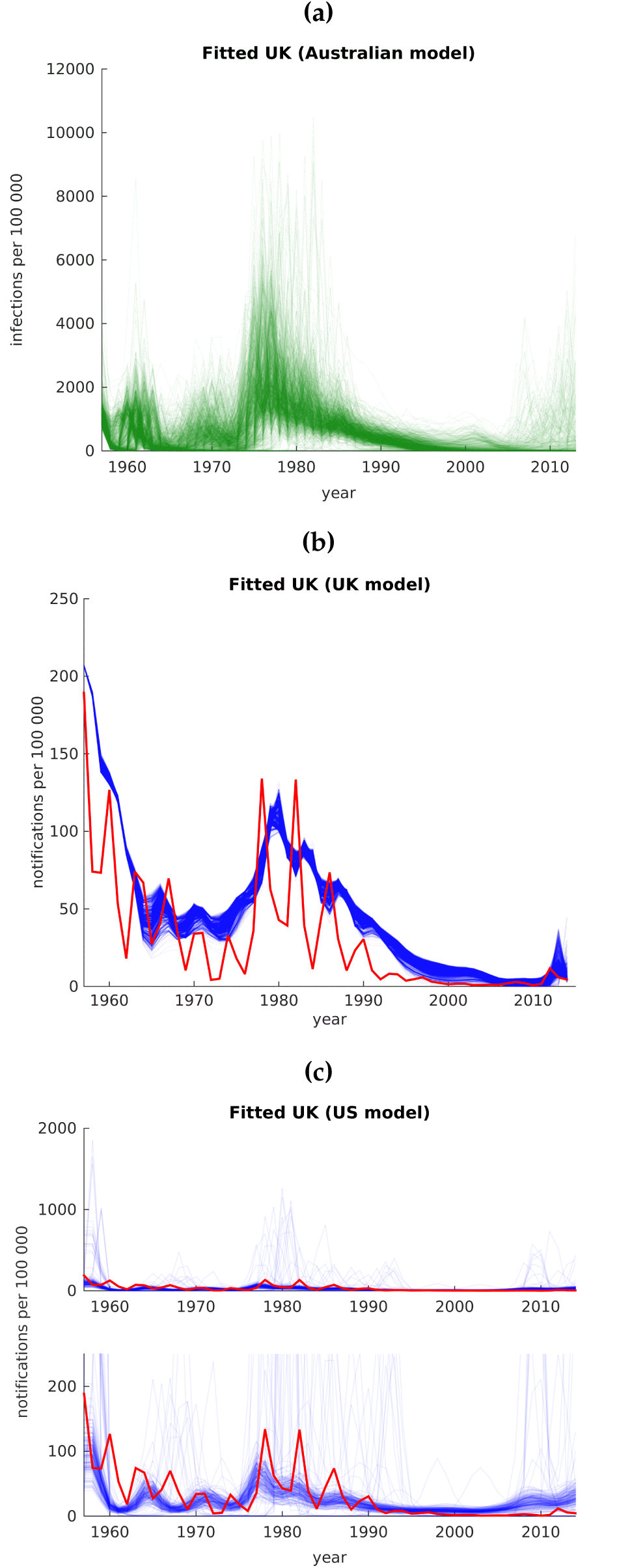
Fitted UK pertussis epidemiology in a) the Australian model, b) the UK model and c) the US model. Note that the Australian model produces infections per 100,000 (shown in green) while the UK and US models produce notifications (shown in blue). In b) and c), the red solid line shows the UK’s published notifications used to fit the model. Part c) shows simulations with the US model using two different scales to include all points — the bottom panel zooms in over the scale of the majority of simulations. Both the UK and the US models closely reproduce the reduction in notifications following the introduction of mass vaccination and the large rebound during the late 1970s/early 1980s. The reduction in infections due to the introduction of vaccination is not observed to the same extent in the Australian model, although the timing and relative magnitude of the resurgence in the late 1970s/early 1980s is consistent with the other two models. All models produce a long period of reduced incidence through the 1990s and early 2000s, but the timing and magnitude of the 2012 epidemic differ between the models — in the Australian model, around 25% of simulations produce resurgence from the mid-2000s; the timing of the 2012 epidemic is reproduced well by the UK model, although the modelled epidemic size is much larger; and the US model produces a gradual increase in notifications following the replacement of whole cell vaccine with acellular vaccine rather than a sharp increase in incidence.

**Figure 7:**
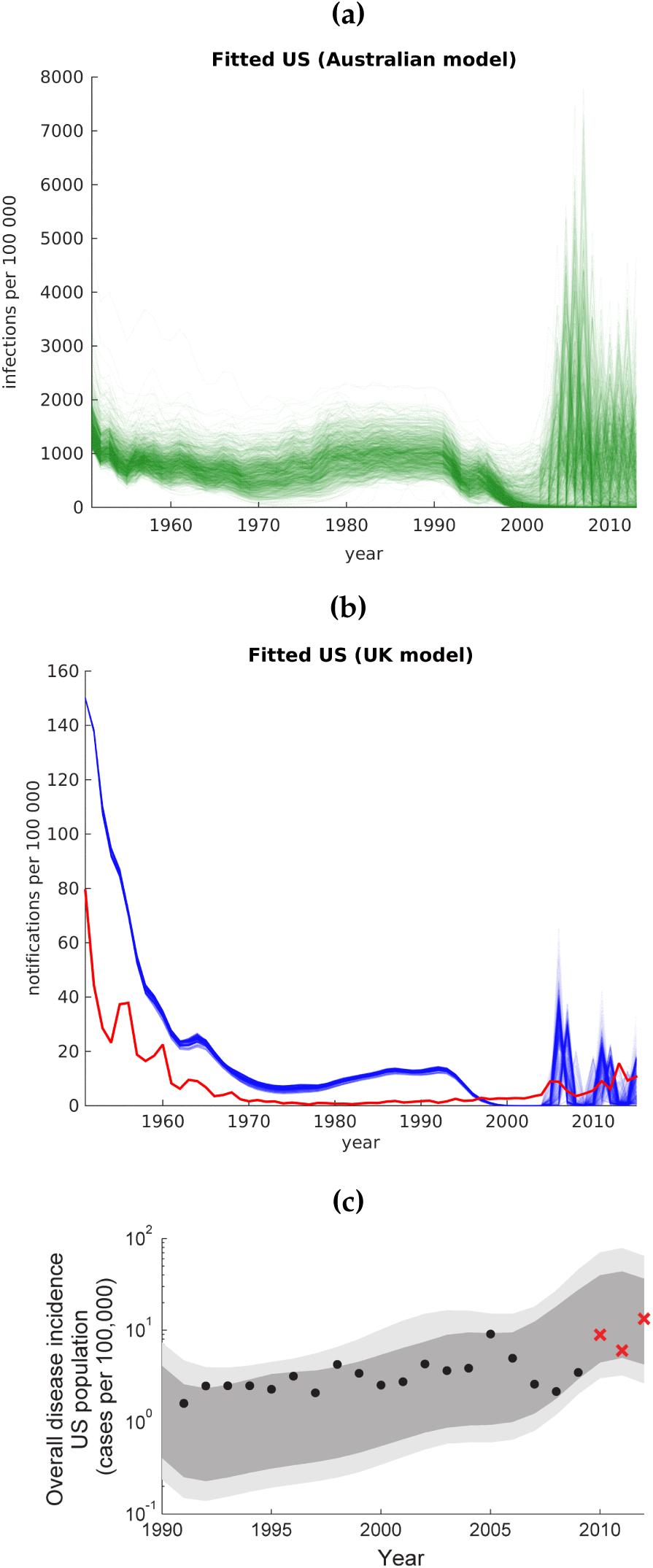
Fitted US pertussis epidemiology in a) the Australian model, b) the UK model and c) the US model. Note that the Australian model produces infections per 100,000 (shown in green) while the UK (shown in blue) and US models produce notifications. In b), the red solid line shows the US published notifications used to fit the model. Note that the US model was not re-fit to US epidemiology during this project and thus the plot is reproduced here from [3] for reference only, noting that it only covers the period from 1990 onwards. Both the Australian and UK models produce a reduction in incidence of infection/notifications when mass vaccination is introduced, although the effect is less marked in the Australian model. A gradual increase in infections/notifications begins in the late 1970s/early 1980s in both models, followed by a fall in the early 1990s. Notably, this reduction in incidence in the early 1990s is produced by all of the models, including the US model (see figure 4c), but is not seen in the observed case data. Both the Australian and UK models generate a return to large epidemics from the mid-2000s, consistent with the timing of resurgence in the US, but of a larger relative magnitude than observed.

#### 4.2.2 Parameter comparison

Best fitting parameters derived using the Australian (figure 8), UK (figure 9) and US (figures 10 and 11) models were compared for each setting. Due to the different model structures, it is important to note that most parameters are not directly comparable across the models. Parameters for vaccine efficacy are only compared between the UK and US models, as the Australian model implemented per-dose efficacy for each of the three primary course doses and booster doses rather than a single value.

**Figure 8:**
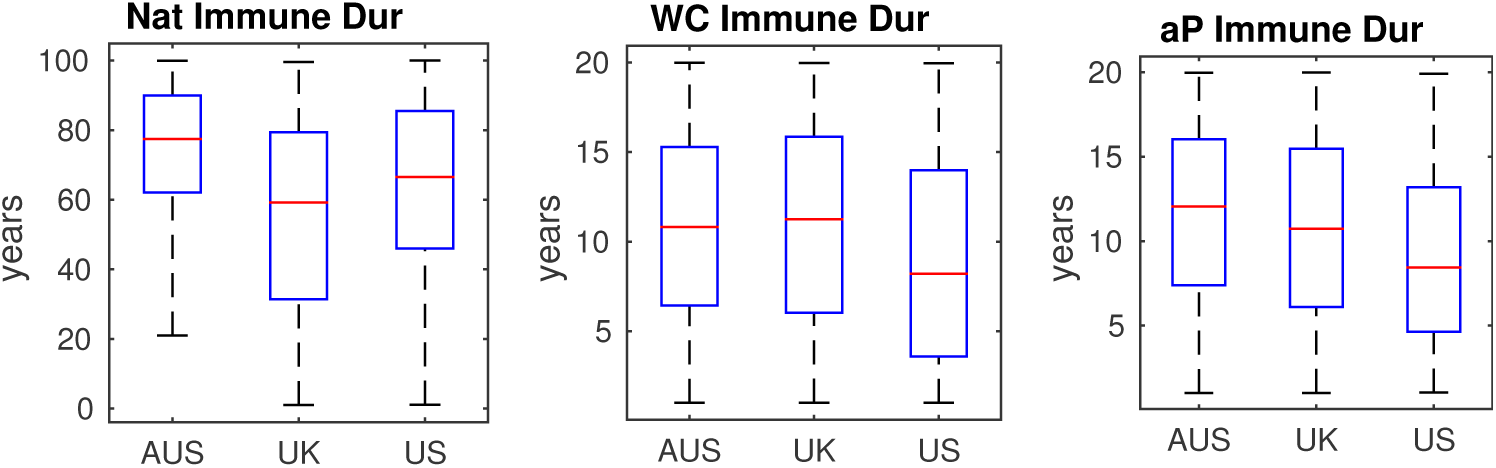
Comparison of selected fitted parameter values for the Australian model. While the duration of natural immunity had median 60–80 years, the duration of vaccine immunity was much shorter.

**Figure 9:**
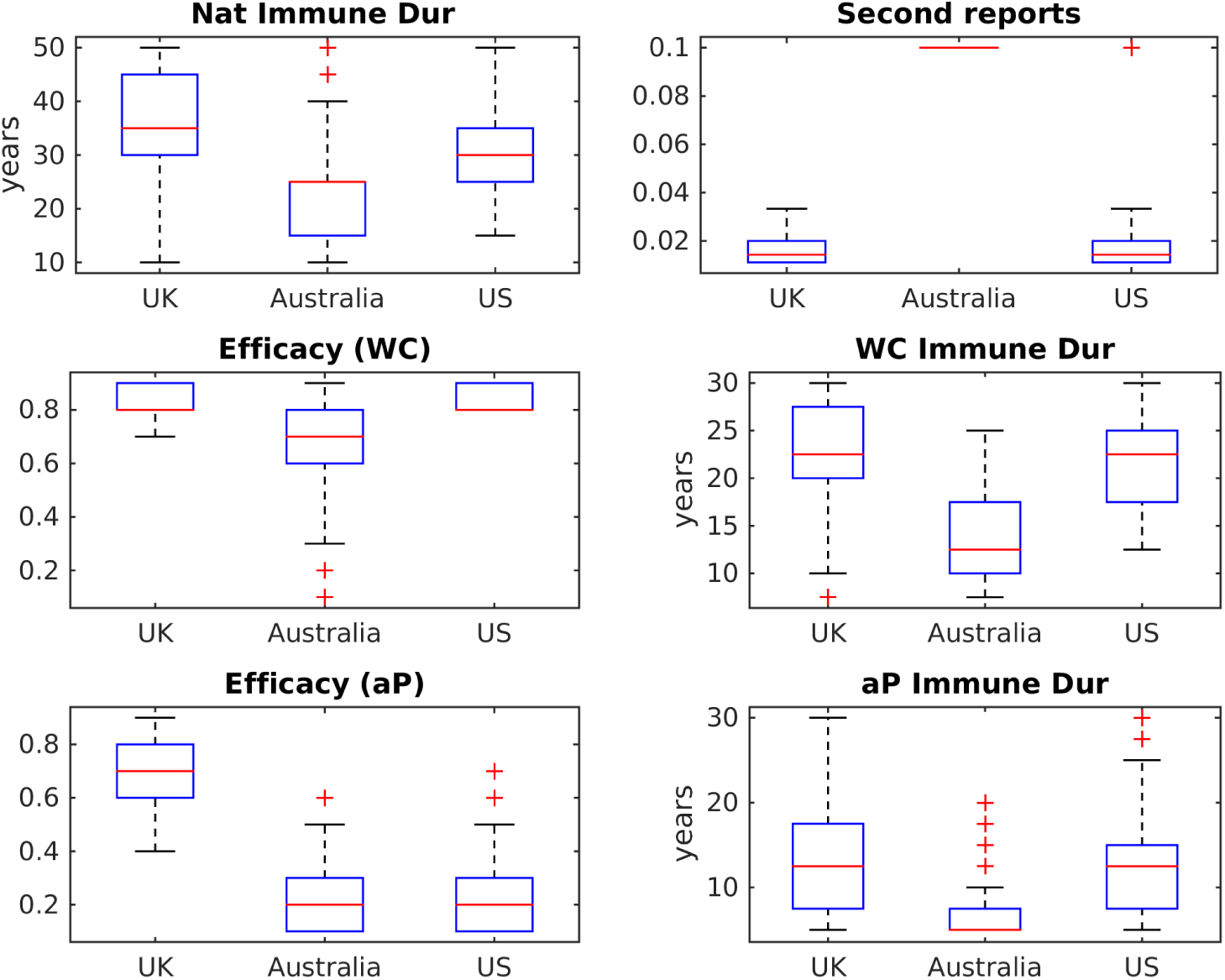
Comparison of all fitted parameter values for the UK model. Fitted parameter values were generally not consistent between countries. Of particular note, selected parameters for Australia tend towards those that would increase the size of the susceptible population to allow more infections to occur, a reflection of the very high notification rates in Australia.

**Figure 10:**
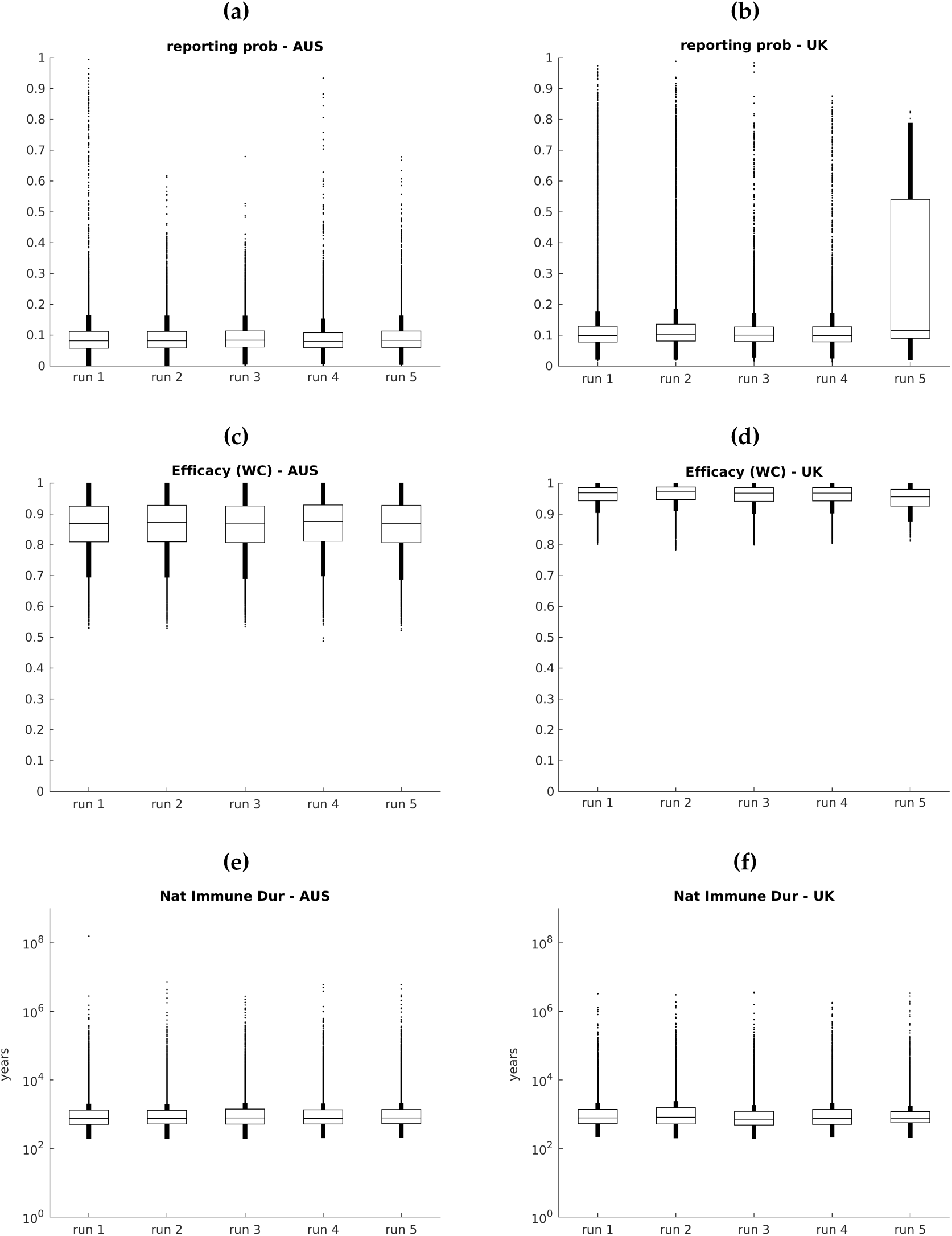
Comparison of selected fitted parameter values for the US model. The left column represents values from fitting to Australia and the right column fitting to the US, for each of the five MCMC runs performed. Posterior parameter distributions were generally consistent across the five MCMC runs for each country. The fitted value for whole cell vaccine efficacy was lower for Australia than for the UK.

**Figure 11:**
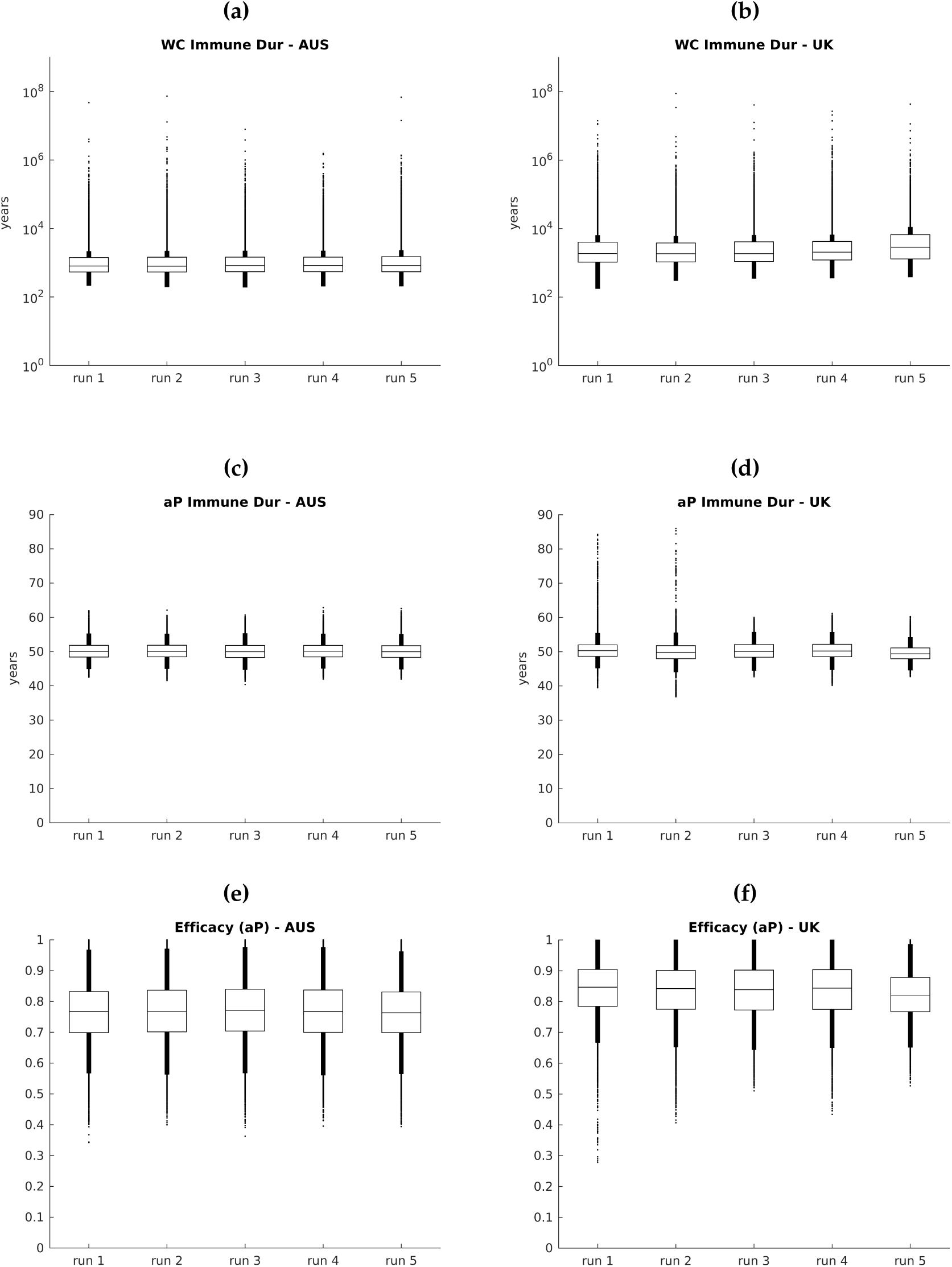
Comparison of fitted parameter values for the US model. The left column represents values from fitting to Australia and the right column fitting to the US, for each of the five MCMC runs performed. Posterior parameter distributions were generally consistent across the five MCMC runs for each country. The fitted value for acellular vaccine efficacy was lower for Australia than for the UK.

For the Australian model (figure 8), only immune duration parameters had similar counterparts in the other models. The duration of immunity following infection in selected simulations was long for each country (with median around 60–80 years), with slightly shorter durations for the UK setting. Duration of immunity following whole cell or acellular vaccination cannot be reliably estimated from the posterior distributions of these parameters, that reflect the distribution from which these values were sampled rather than identifying a clear difference between the two vaccines.

For the UK model (figure 9), selected parameters for each setting showed considerable variability. Parameter combinations selected for Australia generally had very short durations of immunity following infection or vaccination and very low efficacy of acellular vaccine. These values are likely to be a result of the second infection reporting proportion being too low for Australia, and the filtering algorithm selecting those parameter combinations producing the largest possible susceptible population. Sampling the second infection reporting proportion from a wider range would possibly produce values for these parameters more consistent with the UK and US settings, as well as improving model fit.

For the US model (figures 10 and 11), parameter estimates across the five simulation runs for each country were consistent, with the exception of reporting probability in the UK setting. The substantial variability in the range for the reporting probability likely reflects that while the fitting process located a plausible region of parameter space, there may be other suitable regions of parameter space. Some between-country differences can be seen in the vaccine efficacy values for both whole cell and acellular vaccines.

#### 4.2.3 Consistency of results across models and settings

In table 1, incidence and parameter values arising from the refitting process are compared to determine whether the models produce consistent results in different settings, whether the models inform the characteristics of pertussis transmission and immunity and whether the causes of resurgence were consistently identified by each model.

**Table 1:** Evaluation of fitted Australian, UK and US models against key questions examined in the project.

| Key question | Result |
| --- | --- |
| Do the models produce consistent results for a given setting? | **Australia:** General patterns of incidence are similar in all models until the mid-2000s, when the UK (and to a lesser extent the US) model does not reproduce the large 2008–2012 epidemic (figure 5)<br>**UK:** There are some differences in incidence patterns reported by the three models, although all produce increased incidence in the early 1980s followed by a long period of apparent control with some resurgence in the 2000s (figure 6)<br>**US:** Incidence patterns are similar for both the Australian and UK models, although both models produce a downturn in incidence in the early 1990s not seen in the data (figure 7), similar to that produced by the US model (figure 4c) |
| Have the models identified broadly congruent characteristics of pertussis infection and immunity? | **Duration of immunity:** In all models the duration of natural immunity was found to be greater than the duration of acellular vaccine immunity, although the magnitude of natural immunity duration varied substantially across settings. While both the Australian and UK models found the duration of whole cell immunity to be shorter than the duration of natural immunity, the US model found both were essentially lifelong.<br>**Vaccine efficacy (only reported in the UK and US models):** Fitted values of whole cell vaccine efficacy were slightly higher than for acellular vaccines, although values were not consistent between settings. In the UK model, the efficacy of acellular vaccines was particularly low for both Australia and the US at around 20%, but this may reflect the difficulty reproducing very high notification levels in this model.<br>**Probability of case reporting (only reported in the UK and US models):** Fitted probability of case reporting (primary infection only) using the US model was around 8–9 % for both Australia and the UK, somewhat lower than the fitted value of primary infections in using the UK model, which was around 13–14%. |
| Do the models provide similar reasons for pertussis resurgence? | While each model was reasonably able to reproduce resurgence in the country for which it was originally configured, generating resurgence consistent with the magnitude and timing in other countries was more difficult. Without reproducing the epidemiology in each country, it has not been possible to determine reasons for resurgence in each country. |

## 5 Feasibility of a larger study

Each of the three pertussis models has been configured in a particular way to reproduce the observed epidemiology in its original setting. While all models worked well in the setting for which they were designed, some of the assumptions seemed less appropriate when applied to the other two countries. Strong assumptions on the range of values of key pertussis characteristics are likely to have influenced the fitted values for other model parameters, and contributed to the inability of the models to capture all of the epidemiologic features in different countries. This experience is likely to be exacerbated in settings with even less information available for model parameterisation. In addition, applying models to different settings to inform the burden of disease requires assumptions about how pertussis infections relate to pertussis disease in a given setting, shown in this study to be particularly difficult to estimate.

Limited availability of time series incidence data is likely to make fitting especially difficult, similar to the experience in trying to fit the models to the Australian data in this study. The lack of detailed notification data in the pre-vaccination and whole cell eras for this exercise meant assumptions were required about the age distribution of disease in the pre-vaccination era. The impact of these assumptions may well be large in the acellular vaccine era, and additional analyses would be required to determine how sensitive the findings are to assumptions around incomplete data.

Similarly, fitting each of the three models to different settings requires detailed historical vaccination data, with all models displaying sensitivity to changes in coverage. It is important to note that in this study, each model was implemented using the same techniques as were originally used — the results produced here are due to the combination of the model and its implementation, and may not necessarily reflect whether the underlying processes contained in the model equations are an accurate representation of pertussis dynamics.

## Data Availability

All data produced in the present study are available upon reasonable request to the authors

## Appendix

### Results from additional simulation runs of the US model

**Figure 12:**
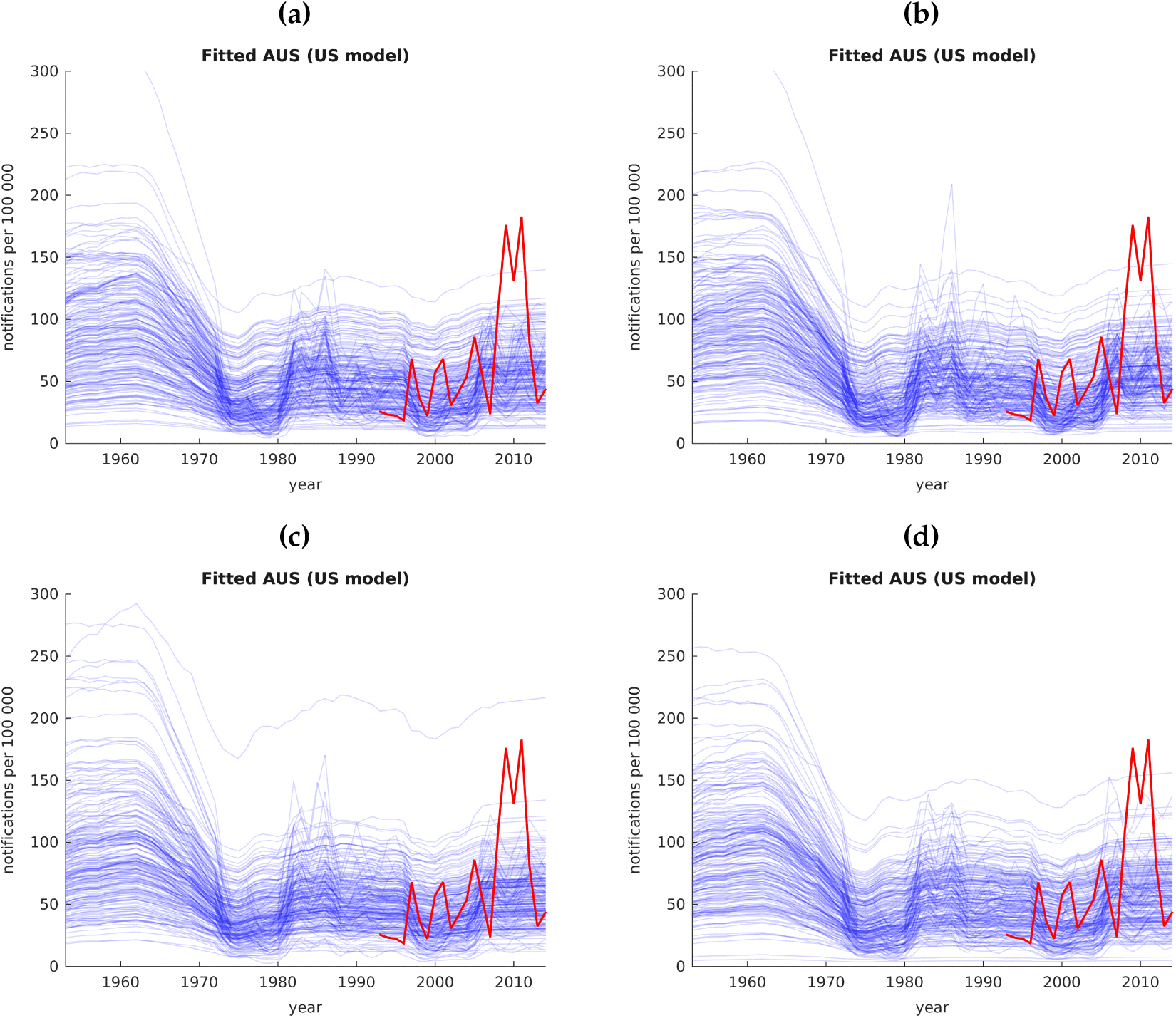
Fitted Australian notifications per 100,000 using the US model. The red solid line shows observed Australian notifications. Five simulation runs of 50,000 each were performed — modelled notifications from an unbiased sample of 200 simulations for the first run are provided in figure 5c, with results for the remaining four runs shown here.

**Figure 13:**
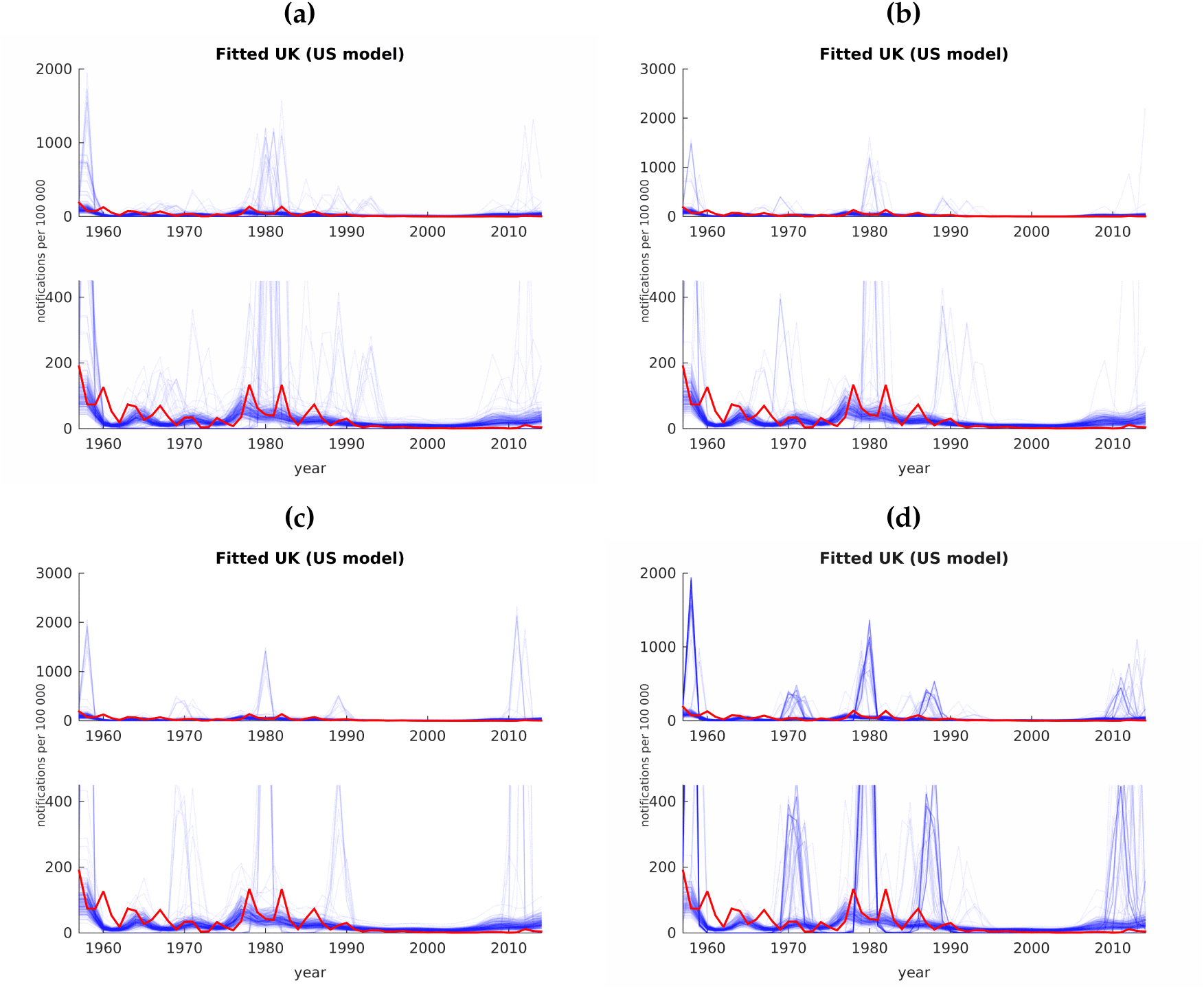
Fitted UK notifications per 100,000 using the US model. The red solid line shows observed UK notifications. Each plot shows simulations with the US model using two different scales to include all points — the bottom panel zooms in over the scale of the majority of simulations. Five simulation runs of 50,000 each were performed — modelled notifications from an unbiased sample of 200 simulations for the first run are provided in figure 6c, with results for the remaining four runs shown here.

